# Loneliness Forecasting Using Multi-modal Wearable and Mobile Sensing in Everyday Settings

**DOI:** 10.1101/2023.06.08.23291165

**Authors:** Zhongqi Yang, Iman Azimi, Salar Jafarlou, Sina Labbaf, Jessica Borelli, Nikil Dutt, Amir M. Rahmani

## Abstract

The adverse effects of loneliness on both physical and mental well-being are profound. Although previous research has utilized mobile sensing techniques to detect mental health issues, few studies have utilized state-of-the-art wearable devices to forecast loneliness and comprehend the physiological manifestations of loneliness and its predictive nature. The primary objective of this study is to examine the feasibility of forecasting loneliness by employing wearable devices, such as smart rings and watches, to monitor early physiological indicators of loneliness. Furthermore, smartphones are employed to capture initial behavioral signs of loneliness. To accomplish this, we employed personalized machine learning techniques, leveraging a comprehensive dataset comprising physiological and behavioral information obtained during our study involving the monitoring of college students. Through the development of personalized models, we achieved a notable accuracy of 0.82 and an F-1 score of 0.82 in forecasting loneliness levels seven days in advance. Additionally, the application of Shapley values facilitated model explainability. The wealth of data provided by this study, coupled with the forecasting methodology employed, possesses the potential to augment interventions and facilitate the early identification of loneliness within populations at risk.

## I. Introduction

Loneliness is a negative feeling caused by loss and disappointment [1]. It arises when individuals assess their existing relationships against their own wishes and societal expectations [2]. Loneliness can have negative impacts on physical and mental health, leading to heightened rates of morbidity and mortality [3]. Multiple studies, such as the research conducted by Park *et al*. [4], have provided compelling evidence establishing a connection between loneliness and adverse physiological and mental health outcomes. Notably, associations have been identified between loneliness and disturbances in sleep patterns, as well as diminished cardiac output. Furthermore, the ongoing COVID-19 pandemic has amplified concerns surrounding loneliness, particularly among adolescents and young adults hailing from lower socioeconomic backgrounds [5]. The profound implications for health underscore the imperative of comprehending the underlying factors contributing to the development of loneliness, as well as the optimal timing for its detection and forecasting.

Existing studies have capitalized on wearable and mobile sensing to detect loneliness. Such works encompass the use of smartphones, wearable sensors, wearable activity trackers, and other technologies to collect information on environmental context, heart rate, activity, and sleep patterns. Wu *et al*. utilize smartphones to collect geosocial data such as the individual’s location and social interaction to detect loneliness in realtime [6]. Li *et al*. capture the app usage information as additional features to identify loneliness risk [7]. Doryab *et al*. employ smartphones to capture behavioral data, including locations, calls log, screen status, etc. as well the FitBit sensors to capture mobility and physical activity to detect loneliness [8]. Besides loneliness, a few studies incorporate mobile sensing to identify other mental health outcomes such as stress, depression, and suicidal ideation [9]–[11].

Nevertheless, current studies predominantly concentrate on the immediate detection of loneliness or offline analysis, failing to account for temporal variations in time series data or explore the interplay between objective data and loneliness. This glaring gap in the literature pertains specifically to the forecasting of loneliness levels with a time interval between the predictive features and the target outcome. More precisely, there exists a lack of research pertaining to the forecasting of loneliness over a defined period, leading to an insufficient availability of dedicated datasets for addressing this specific research inquiry.

Modern wearable devices (such as the Oura Ring and Samsung smartwatch) and mobile phones have demonstrated their capacity to assess a wide range of physiological, behavioral and contextual information. Their viability for integration into healthcare applications has been substantiated by previous research [12], [13]. The incorporation of such supplementary information holds promise as a prospective avenue for exploration in the modeling of loneliness. Furthermore, with regard to the intricate relationship between loneliness and other health risks, the potential forecasting or early detection of feelings of loneliness presents novel opportunities for the development of targeted interventions aimed at alleviating loneliness and enhancing individual well-being. By leveraging the ubiquitous sensing capabilities of modernized devices, it becomes possible to continuously gather an extensive array of physiological and behavioral data in tandem with self-reported levels of loneliness to build models.

In this paper, we present a personalized machine-learning method to forecast levels of loneliness by leveraging longitudinal data encompassing physiological, behavioral, and contextual information collected from smart rings, watches, and smartphones. To achieve this, we conducted a two-month monitoring study involving a cohort of 30 college students, utilizing the Oura Ring, Samsung Smartwatch, and the AWARE app installed on smartphones. In addition to capturing objective and passive data through these devices, we collected self-reported loneliness information from the participants, thereby generating detailed and granular labels for evaluation purposes. The collected objective and subjective data were employed to train and assess the effectiveness of our proposed loneliness forecasting method. Moreover, we integrated SHAP (SHapley Additive exPlanations) to compute the Shapley values, allowing for a deeper understanding of the relationship between the temporal features derived from these time series data and the experience of loneliness [14].

## II. Loneliness Data Collection

Our study encompasses a comprehensive approach to data collection, encompassing self-reported loneliness alongside associated physiological and behavioral information. To facilitate efficient data collection and storage, we implemented a robust platform. The data obtained during the study can be categorized into three distinct types: objective physiological data, objective behavioral data, and subjective questionnaires. Objective physiological data were acquired utilizing the Oura ring and Samsung smartwatch. The Oura ring enabled the assessment of sleep patterns, while the Samsung smartwatch facilitated continuous monitoring of physiological parameters throughout the day. Objective behavioral data were collected through the implementation of the AWARE [15] phone app, which captured mobile sensory data and recorded detailed information regarding participants’ phone usage patterns. Subjective self-report data were gathered employing our custom mSavorUs [16] phone app, specifically designed for the collection of subjective experiential data and the provision of interventions. The self-reported loneliness levels were measured on a scale ranging from 0 to 100, with 0 indicating “not feeling lonely at all” and 100 indicating “feeling extremely lonely.” These self-reported loneliness levels were further categorized as either “Not Feel Lonely” or “Feel Lonely,” based on whether they fell below or above the population median, respectively. All procedures were approved by the researchers’ academic institutional review board (#2019-5153).

### A. Participants

This study focuses on examining the relationship between loneliness and mental health within a cohort of 30 full-time college students enrolled at a university located in Southern California. The selection criteria for participants ensured a homogeneous sample by excluding married students, individuals with children, those returning to school after a three-year hiatus, and those with severe psychopathology. Eligible participants were required to possess fluent English language skills and an Android smartphone compatible with both the Oura Ring and Samsung Active 2 watch, which were utilized for data collection purposes. The recruitment process involved outreach to faculty members, dissemination of information through social media platforms, and the implementation of screening surveys to identify potential participants with symptoms of depression or suicidal ideation.

### B. Recruitment

Participants meeting the inclusion criteria underwent a baseline assessment in a lab session and set up the devices under instruction. During the monitoring phase for approximately 8 weeks, participants are asked to wear the devices continuously, except during charging or potentially damaging activities. The watch app collected 12 minutes of photoplethysmogram (PPG) signals every 2 hours. The AWARE app monitored phone usage, while the mSavorUs app prompted participants for brief surveys multiple times daily.

## III. Method

In this study, we devise a loneliness forecasting model on predicting loneliness seven days in advance. Given the complex nature of the multi-modal time series data, we employ machine learning models to perform feature extraction and classify the levels of loneliness. Our proposed methodology encompasses the key components including the extraction of physiological, behavioral, social, and contextual features. We address challenges such as feature alignment, handling missing data, and the development of a classification model.

### A. Feature Extraction

#### 1) Physiological Features

We extract HR and HRV-related features from the signal collected from smart rings and watches as the physiological features. In ubiquitous monitoring settings, signal collection with PPG-based wearable devices can be affected by the presence of noise. To overcome this issue, we implement the methods to extract Heart Rate (HR) and Heart Rate Variability (HRV) features from raw collected PPG signals [17]–[19]. We first conduct a signal quality assessment to classify PPG signals as clean or noisy, then reconstruct short-term noisy segments via a generative adversarial network. Then the systolic peaks and inter-beat intervals are detected by a dilated Convolution Neural Network, from which HR and HRV-related features are extracted.

#### 2) Behavioral Features

Behavioral features refer to the patterns and characteristics of subjects’ smartphone usage behavior. They are extracted from participants’ smartphone usage. We extract the smartphone usage features, including the number of battery charger plugins, screen off, screen on, screen locks, and screen unlocks.

#### 3) Social Features

The term “social features” pertains to the discernible patterns of social activity demonstrated by individuals through their smartphone usage. These features serve as valuable indicators that shed light on the nature of individuals’ interactions and engagements within their respective social networks. Examples of such social features encompass the quantity of messages and notifications within various categories, as well as the duration and frequency of phone calls within specific time windows.

#### 4) Contextual Features

Contextual features encompass the environmental information related to the subjects. These features provide insights into the surrounding context in which individuals operate using GPS localization. The contextual features include the variance of latitude, variance and mean of speed, number of places, duration at home, the mean and standard deviation of the duration outside, and the total travel distance within a given location time window.

### B. Feature Alignment

The dataset contains features collected at varying resolutions, with some features recorded once daily (e.g., sleep-related) and others, such as PPG-related features, captured multiple times a day. To align the features with each loneliness label, we average the values within a designated time window preceding the corresponding loneliness levels. The selection of the optimal time window for each feature is based on the averaged feature’s correlation with the target loneliness levels. Subsequently, the resulting data records were compiled using the aligned features corresponding to the optimal window lengths and the self-reported loneliness level.

### C. Missing Data

In our dataset, missing data may occur due to participants forgetting to wear or charge their devices. We discovered that using the average of all valid values for each participant exhibited a strong correlation with the target questionnaire responses for most features. Therefore, we implement a single imputation method to address missing data across all features [20].

### D. Classification Model

We define a binary classification task to forecast loneliness. Each self-reported loneliness is associated with the aligned features within a 14-day time window, starting 3 weeks prior to the loneliness target. There is a one-week interval between the available features and the target, indicating a forecasting task. However, the number of aligned feature records within the time window may vary due to different self-report frequencies. To address this, we aggregate the data by calculating the daily average for each aligned feature. This aggregation process ensures that the number of features for each loneliness target remains consistent, corresponding to the number of time window length times the daily feature amount. To forecast loneliness with the aggregated features, we employ Random Forests consisting of 400 trees with a maximum depth of 15.

### E. Model Explainability

We explore the model explainability by utilizing path-dependent feature perturbation algorithms [21] to compute the Shapley values of the features. Based on the Shapley values for each feature, we obtain a quantitative measure of their contributions to the predictions. These values allow us to rank the features based on their relative importance and assess their effect on the output loneliness category.

## IV. Results

### A. Loneliness Forecasting Performance

The models are trained and validated in a personalized manner on the data from a two-month study involving 30 participants who completed multiple self-report assessments each day. Each participant’s data were divided into a test set and a train set. The test set contains the most recent 50% of the data in the monitoring. However, the training set includes the first 50% data of the same person as well as data from all other participants. Personalized models were then trained and tested using these training-testing set pairs from each participant. This approach allows us to assess the model’s performance by simulating real-world scenarios while maintaining the integrity of individual participant data.

After preprocessing, we obtained a total of 6212 data points, which were used to develop 30 personalized models to forecast loneliness 7 days ahead using the features from the past two weeks. To assess the performance of our approach, we average the results across all the personal models. The overall performance is summarized in Table I. Our loneliness forecasting model achieves an accuracy of 0.823, recall of 0.905, precision of 0.750, and F-1 score of 0.820. In addition, our model achieves Cohen’s kappa of 0.648, indicating substantial agreement between the predicted and true loneliness.

**TABLE I.** The overall performance on loneliness forecasting.

| Accuracy | Recall | Precision | F-1 | Cohen's Kappa |
| --- | --- | --- | --- | --- |
| 0.823 | 0.905 | 0.750 | 0.820 | 0.648 |

Table II illustrates the confusion matrix of the loneliness detection models, derived from 3095 test samples. This suggests that, based on the physiological or behavioral, social, contextual information, the model could forecast if the participant would feel lonely. Note that the overall performance of our model is influenced by false negatives.

**TABLE II.**
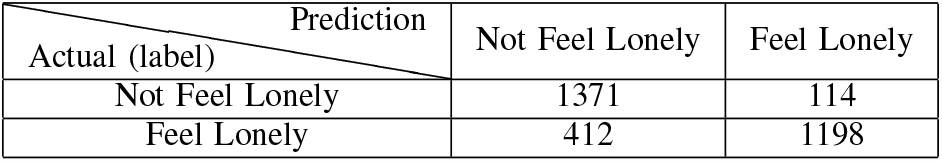
Confusion matrix

### B. Model Explainability

Figure 1 presents the overall Shapley value across all test samples for the top 20 features. The feature names are noted with the prefix ‘day*x*’ indicating the specific day within the time window from which each feature originates. Each dot in the plot represents a feature value instance, with its horizontal position indicating its effect on the predicted loneliness category. Dots positioned to the left suggest a higher probability of predicting “Not Feel Lonely” while dots positioned to the right indicate a higher probability of predicting “Feel Lonely”.

**Fig. 1.**
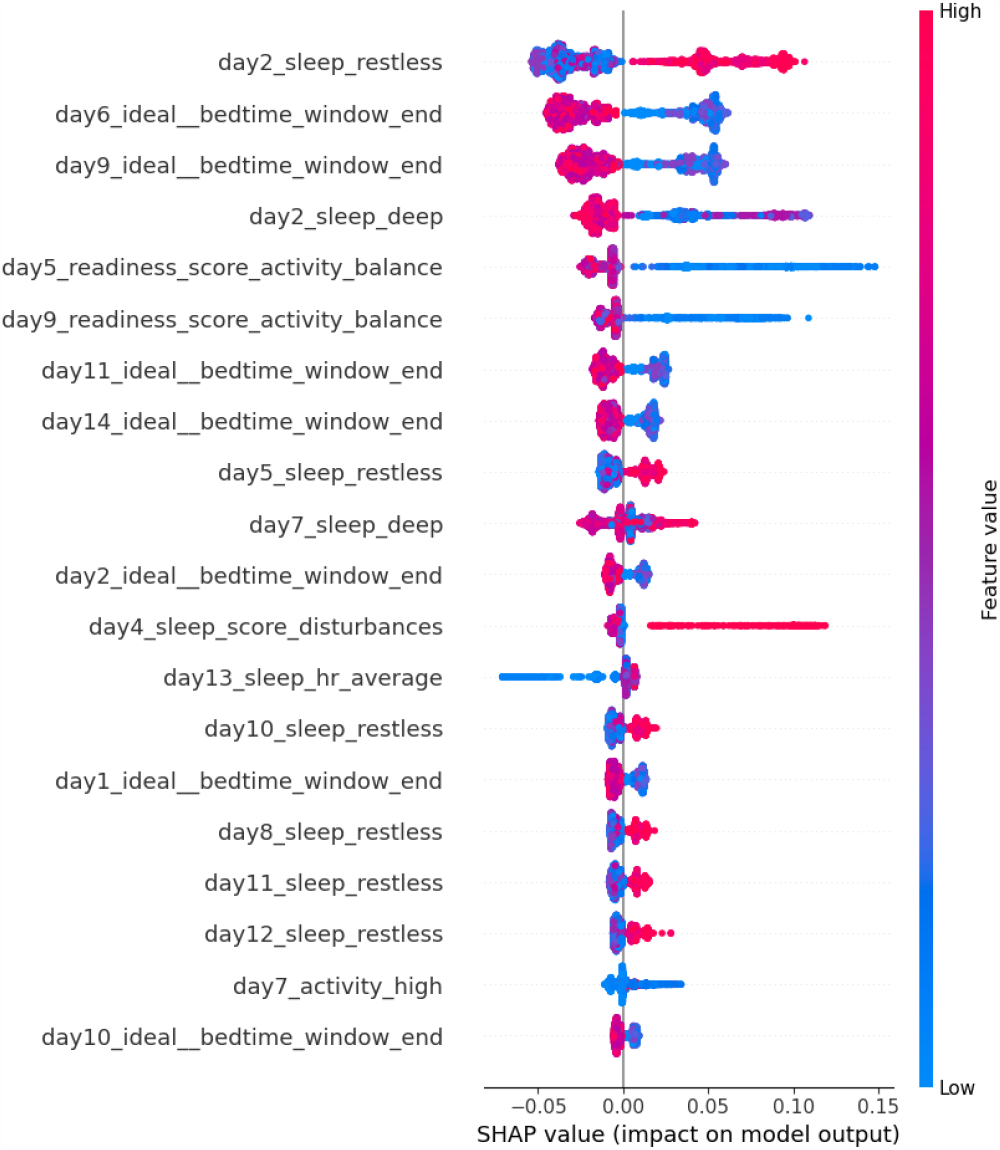
Feature Importance Using Shapley Values Beeswarm Plot

We observe that higher sleep restless scores are strongly associated with increased loneliness likelihood, making it the most influential feature. Lower activity balance scores also contribute significantly, implying that lower activity levels are associated with an increased likelihood of feeling lonely.

## V. Conclusion

This paper proposed a machine learning method for forecasting loneliness based on physiological, behavioral, and contextual information collected from smart rings, watches, and smartphones Our developed model achieved an accuracy of 0.823 and an F-1 score of 0.820 in forecasting loneliness levels 7 days ahead. We incorporated Shapley values as explainability methods to gain insights into the relationship between the features and loneliness. By extending beyond existing studies that focus on prompt loneliness detection, our approach offered the potential for clinical implementation in the early identification and addressing of loneliness among at-risk populations. Further investigation into the utilization of deep learning models for predicting loneliness has the potential to improve performance by automatically extracting more robust features. Moreover, deep learning holds promise in forecasting loneliness in several weeks.

## Data Availability

All data produced in the present study are available upon reasonable request to the authors

